# Dissecting PON1 Genotype Combinations Modulating Paraoxonase Activity and Risk of Dysglycemia and Liver Fibrosis

**DOI:** 10.64898/2026.04.09.26350501

**Authors:** Laura Herrera, Maria João Meneses, Rogério T. Ribeiro, Luís Gardete-Correia, João F. Raposo, José Manuel Boavida, Carlos Penha-Gonçalves, M. Paula Macedo

## Abstract

**Background & Aims:** Metabolic disorders such as dyslipidemia, metabolic dysfunction–associated steatotic liver disease (MASLD), and diabetes are promoted by chronic pro-inflammatory and pro-oxidative states. Paraoxonase 1 (PON1), a liver-derived HDL-associated enzyme, plays an important antioxidant role by hydrolyzing oxidized lipids and protecting against oxidative stress– induced damage. Genetic variation in PON1, particularly in promoter and coding regions, modulates enzyme expression and activity, thereby influencing susceptibility to metabolic and cardiovascular diseases. This study investigated the genetic determinants of serum paraoxonase (PONase) activity and their relationship with dysmetabolic phenotypes.

**Methods:** A genome-wide association study was conducted in 922 Portuguese individuals from the PREVADIAB2 cohort. Genetic variants and haplotypes related to PONase activity were analyzed, and associations with dysglycemia and liver fibrosis were evaluated in individuals aged over 55 years.

**Results:** We identified two key PON1 variants as determinants of PONase activity: rs2057681 (in strong linkage disequilibrium with the non-synonymous Q192R variant) and rs854572 (located in the promoter region). Analysis of rs854572–rs2057681 haplotypes revealed that specific combinations differentially modulate PONase activity and confer risk or protection for dysglycemia and liver fibrosis, depending on the rs2057681 genotype context. Notably, although PONase activity was strongly associated with PON1 variants, it did not directly correlate with dysmetabolic phenotypes, suggesting that genetic context and haplotype structure, rather than enzyme activity alone, shape disease susceptibility.

**Conclusions:** These findings highlight the complex genetic architecture of PON1 and its role in metabolic disease risk, supporting the use of PON1 genetic information to uncover predisposition to dysmetabolic conditions. Our results provide insights into the interplay between PON1 genetics, enzyme function, and dysmetabolism, with implications for risk stratification in metabolic liver disease.

**Lay Summary:** PON1 is a liver-derived gene that encodes an enzyme involved in protection against oxidative stress, a key contributor to metabolic liver disease and diabetes. In this study, we found that specific combinations of PON1 genetic variants are associated with abnormalities in blood glucose regulation and with markers of liver fibrosis. These associations were dependent on genetic configuration rather than enzyme activity alone, suggesting that PON1 genetic information may help identify individuals at higher risk of metabolic liver disease.

## INTRODUCTION

Metabolic imbalances of complex and multifactorial nature underpin the interlinking of clinical conditions such as dyslipidemia, metabolic dysfunction-associated steatotic liver disease (MASLD), and diabetes^1,2^. Experimental, mechanistic, and clinical studies strongly support the concept that a chronic pro-inflammatory and pro-oxidative milieu is a central driver of disease progression. In particular, oxidative stress promotes lipid peroxidation, especially of unsaturated fatty acids within cell membranes and lipoproteins, leading to the formation of bioactive oxidized lipid species (e.g., oxysterols, isoprostanes) ^3^. These oxidized lipids disrupt normal lipid metabolism, increasing mitochondrial oxidative stress and sustaining chronic inflammation ^4^. Furthermore, pro-oxidant conditions also impair mitochondrial function, reducing fatty acid oxidation and promoting lipid accumulation (steatosis) in organs such as the liver and kidney ^5,6^. Concurrently, oxidative stress damages cellular components, including proteins and enzymes involved in glucose uptake and metabolism, ultimately impairing insulin signaling and promoting glucose intolerance ^7^.

Paraoxonase 1 (PON1) is an esterase/lactonase predominantly synthesized in the liver and secreted into the bloodstream, where it associates with high-density lipoprotein (HDL) particles^8^. PON1 has a broad substrate specificity, including organophosphates, aryl esters, and, most notably, oxidized lipids such as lipid hydroperoxides and specific lactones generated under oxidative stress^9^. PON1^−^/^−^ mice show increased levels of oxidized lipids and systemic oxidative stress markers, confirming the role of PON1 enzymatic activity in hydrolyzing lipid peroxides and protecting lipoproteins from oxidative modification ^10^.

Accumulating evidence supports a cryptic role for PON1 in the pathophysiology of several oxidative stress-related diseases, particularly atherosclerosis, metabolic syndrome, and MASLD ^11–14^. PON1 anti-oxidant capacity, primarily through its hydrolysis of oxidized phospholipids and lipid peroxides on low-density lipoproteins (LDL), directly mitigates oxidative modification of lipoproteins, a key step in atherogenesis ^13,15,16^. Clinical studies have consistently demonstrated that reduced serum PON1 activity is associated with increased atherosclerosis risk in individuals with familial hypercholesterolemia and in type 1 diabetes, independent of traditional lipid parameters impairments ^11,13,14,16^. Furthermore, low PON1 activity has been associated with increased carotid intima-media thickness and poor vascular endothelial function, underscoring its prognostic value in cardiovascular disease ^11,13,16,17^. In hepatic pathology, reduced PON1 activity is correlated with steatosis, inflammation, and fibrosis in MASLD, suggesting that impaired detoxification of lipid peroxidation products may exacerbate liver injury ^12,14^. PON1 deficiency increases susceptibility to hepatotoxicity and liver steatosis, particularly in high-fat, high-cholesterol– fed mice ^12^, underscoring its role in maintaining hepatic redox balance and lipid homeostasis. Beyond lipid metabolism, PON1 is implicated in modulating inflammation and maintaining endothelial function through its capacity to degrade lipid peroxides and inflammatory mediators ^13,18^. PON1^−^/^−^ mice often exhibit exaggerated inflammatory cytokine responses and endothelial dysfunction, highlighting PON1 influence in immunomodulation and vascular protection ^13,15,18^.

The variation of PON1 expression and activity is under strong genetic control, primarily governed by functional polymorphisms within the PON1 gene. Among the most extensively studied are the promoter region variant −108C>T (rs705379) and the coding region variants Q192R (rs662) and L55M (rs854560) ^19–21^. The −108C allele has been associated with higher transcriptional activity and increased serum PON1 levels, presumably via enhanced affinity for transcription factor binding sites ^20,21^. Similarly, the Q192R and L55M variants modulate enzymatic activity and stability, with the 192Q isoform displaying greater degradation efficacy of lipid peroxides and the 55L variant showing higher plasma protein concentrations ^21^. Additional promoter polymorphisms, such as −832G>A and −1741G>A, though less frequently annotated in public databases, have also been linked to increased PON1 expression and hydrolytic activity, particularly in response to lipid substrates and drugs such as statins ^19,21^.

These genetic differences have clinical relevance, and recent research has reinforced the significant contribution of PON1 gene polymorphisms to the susceptibility and progression of several complex diseases, particularly cardiovascular disorders (CVD), MASLD and diabetes ^20,22,23^. The 192R allele, although more efficient in hydrolyzing certain substrates like paraoxon, is less effective against lipid peroxides, correlating with increased oxidative stress and heightened cardiovascular risk ^19,22^. Promoter variants have been shown to modulate gene expression, with the −108T allele (rs705379) being associated with lower transcriptional activity and serum enzyme levels, contributing to enhanced atherogenic potential and hepatic lipid accumulation ^20,21^. Moreover, recent studies have identified associations between rare PON1 variants and features of MASLD, including liver fibrosis and insulin resistance, enlarging the genetic etiology spectrum of these metabolic dysfunctions ^19,20,23^.

Overall, PON1 serum enzymatic activity represents a mediator of disease progression in lipid-associated disorders but it remains unexplored whether PON1 genetic variants can be instrumental in constructing clinical indicators of dysmetabolism susceptibility. Current findings underscore the complex control of *PON1* genetic variation in modulating enzyme function and influencing the pathogenesis of disorders fueled by oxidative stress. Herein, we hypothesized that specific PON1 genotype configurations controlling serum PON1 activity can be useful in uncovering genetic propensity to develop dysmetabolic conditions such as type 2 diabetes and liver fibrosis.

## Materials and Methods

### Ethical permits and study cohort

All participants were volunteers and provided written informed consent for inclusion in the PREVADIAB2 study ^24^. The protocol followed the principles of the Declaration of Helsinki and received approval from the Ethics Committee of Associação Protectora dos Diabéticos de Portugal (APDP Diabetes Portugal (900/2013)) and NOVA Medical School (CEFCM/02/2020), as well as from the Portuguese Data Protection Authority (permit no. 3228/2013). The study population was drawn from the PREVADIAB2 and initially comprised 924 genotyped subjects.

### Biochemical and Clinical assessments

Anthropometric measurements included body weight (to the nearest 0.01 kg), height (to the nearest 0.1 cm, using a stadiometer), and waist circumference (measured midway between the lowest rib and iliac crest), following WHO guidelines. Determination of glycemic status was based on glucose measurements in blood samples collected after a 12-hour overnight fast and during a standard 75 g oral glucose tolerance test (OGTT) at 0, 30, and 120 minutes. Plasma glucose was quantified by the glucose oxidase method (Olympus AU640, Beckman Coulter). Glycemic status was defined according to WHO/International Diabetes Federation (IDF) criteria ^25^. For analysis purposes, the study population was grouped into normoglycemic (n= 638) or dysglycemic subjects (n= 261, including prediabetes and diabetes).

The Fibrotic NASH Index (FNI) scoring tool ^26,27^ was applied to estimate the probability of fibrotic metabolic dysfunction-associated steatohepatitis (MASH) based on serum measurements of spartate aminotransferase (AST), high-density lipoprotein (HDL) cholesterol using enzymatic methods in an automated analyzer (Olympus AU640, Beckman Coulter) and glycated hemoglobin (HbA1c) measured by high-performance liquid chromatography with boronate affinity (Menarini Premier Hb 9210). For analysis purposes the study population was grouped into individuals with low probability of fibrotic MASH (FNI≤0.1, n=525) and higher probability of fibrotic MASH (FNI>0.1, n= 394).

### PON1 activity analysis

Following Batuca et al. ^28^, serum paraoxonase (PONase) activity was assessed using a substrate mixture containing paraoxon (1.0 mM; Sigma-Aldrich) freshly prepared in 50 mM glycine buffer containing 1 mM calcium chloride (pH 10.5). For each sample, 10 μL of serum were added to 290 μL of substrate mixture and incubated at 37 °C in 96-well plates. The release of *p*-nitrophenol was measured at 412 nm between 10 and 50 minutes of incubation and activity was expressed as units per ml serum.

### Genotyping and data preparation

Genomic DNA was genotyped using the Axiom™ HGCoV2_1 array (Thermo Fisher Scientific). Genotyping was performed by the Microarray Research Services Laboratory, ThermoFisher Scientific, Santa Clara, CA. SNP positions were aligned to the GRCh38/hg38 reference genome, and variant annotation was based on dbSNP v151, the 1000 Genomes Project Phase 3 (2014), and Affymetrix internal screening data (NetAffx build 36, revision r2.a1, July 15, 2020).

### Genome-wide analysis

The initial dataset comprised 832,157 variants in 924 individuals. Standard quality control (QC) filters were applied using PLINK v1.9 ^29,30^ including exclusion of individuals and variants with >5% missing data, variants with minor allele frequency (MAF) <10%, and variants deviating from Hardy–Weinberg equilibrium (*P* < 1×10^−7^). After QC, the dataset included 293,781 variants prior to linkage disequilibrium (LD) pruning. Following LD pruning, the final dataset for genome-wide analysis comprised 240,834 variants and 922 subjects. Genome-wide analysis using the additive and dominant genetic models was performed to test associations of serum PONase activity with SNPs in the pruned dataset. Covariates included age, sex, and body mass index (BMI). Analysis of PON1, PON2 and PON3 gene region using the non-pruned variant set captured 27 SNPs in the chromosome 7 region 95,271,366 bp to 95,449,748 bp, using both additive and dominant models. Stratified analysis for glycemia status (normoglycemia or dysglycemia) and for liver fibrosis risk (FNI ≤0.1or FNI >0.1) was performed in this region to ascertain whether significant genetic associations were modified by dysmetabolic states. To identify independent signals GWAS conditional using rs2057681 genotype as a covariate. Genome-wide significance was defined using Bonferroni correction (p < 0.05/N, where N is the number of SNPs tested) and genomic inflation was assessed using the λ statistic. We report nominal and adjusted *P* values of association tests. Manhattan and QQ plots were generated to visualize genome-wide associations for all analyses using Python Qmplot ^31^and Haploview v4.2 ^32^ was used to construct and visualize linkage disequilibrium (LD) mapping. Haplotype analysis was performed by phasing genotypes with Beagle v5.5 ^33^ focusing on the two SNPs (rs2057681 and rs854572).

### Statistical Analysis

Group comparisons were conducted using the Kruskal–Wallis test with Dunn’s correction performed in GraphPad Prism version 10.2. Relative risks were assessed using Fisher’s exact test for P values, and 95% confidence intervals were computed with the Koopman asymptotic score method. For analyses of dysglycemia and liver fibrosis, participants were restricted to individuals aged over 55 years, an age group in which metabolic dysregulation and fibrotic liver changes are more prevalent and clinically established. This restriction was applied to reduce heterogeneity related to early or transient metabolic alterations.

## Results

### Genome-wide association analysis of serum PON1 activity

Genome-wide association analysis (GWAS) was conducted to elucidate the genetic determinants of serum paraoxonase (PONase) activity in a Portuguese cohort (PREVADIAB2 study) ^24^. After QC procedures the final dataset for GWAS analyses consisted of approximately 240,000 single nucleotide polymorphisms (SNPs) genotyped in 922 individuals that had serum PONase activity quantified. After adjustment for age, sex, and body mass index, a major locus on chromosome 7 emerged as the principal genetic determinant of serum PONase activity (Figure 1A and B). Seven SNPs spanning a 22.2 kb region within the PON1 gene achieved genome-wide significance under a dominant genetic model, following stringent correction for multiple testing (Table 1). These SNPs, which are in strong linkage disequilibrium (LD), define a distinct LD block within the PON1 locus, in the context of 27 genotyped SNPs in the chromosome 7 encompassing PON1, PON2, and PON3 gene regions (Supplementary Figure 1). These findings corroborate previous reports that PON1 genetic variants are major determinants of serum PON1 enzymatic activity ^34^.

**Table 1.**
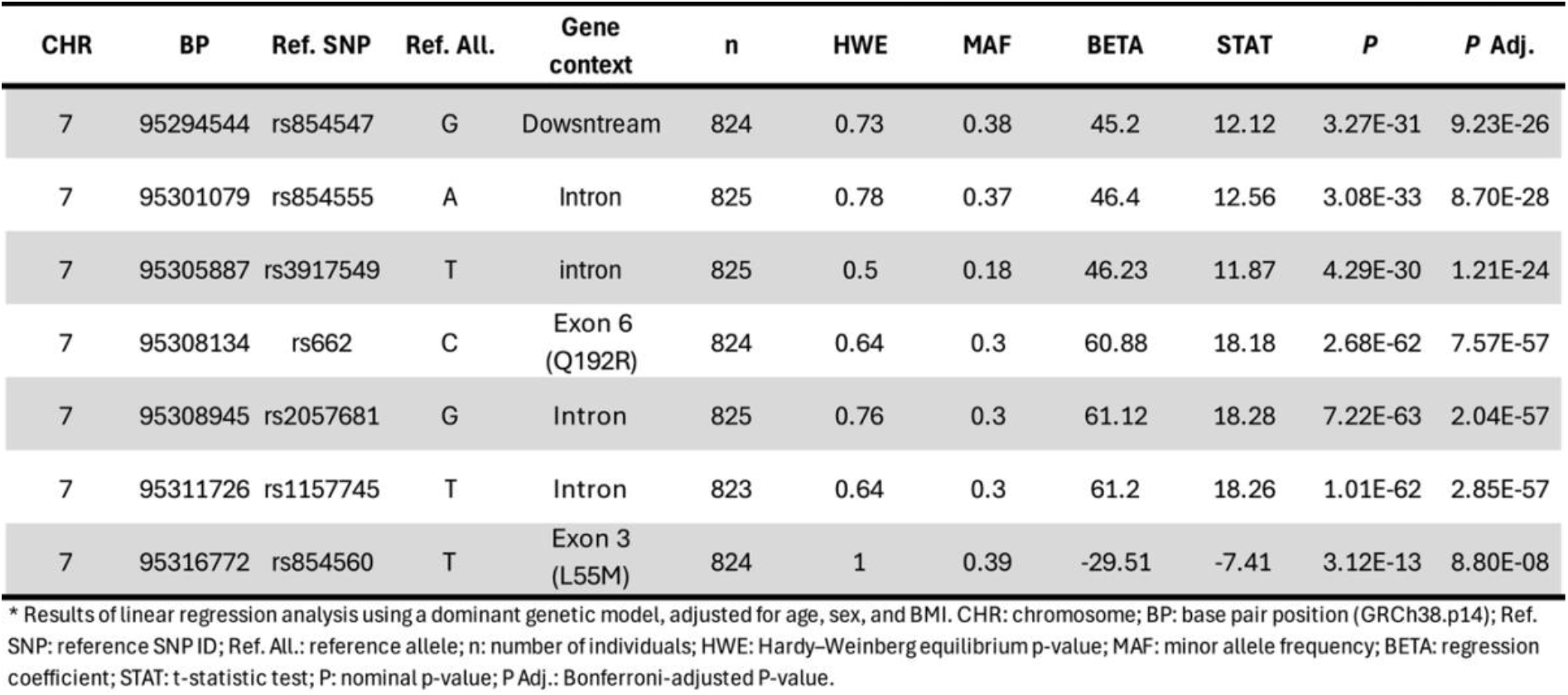
Genome-wide association to PONase serum activity. Seven SNPs showing significant association map within the PON1 gene region^*^.

**Figure 1:**
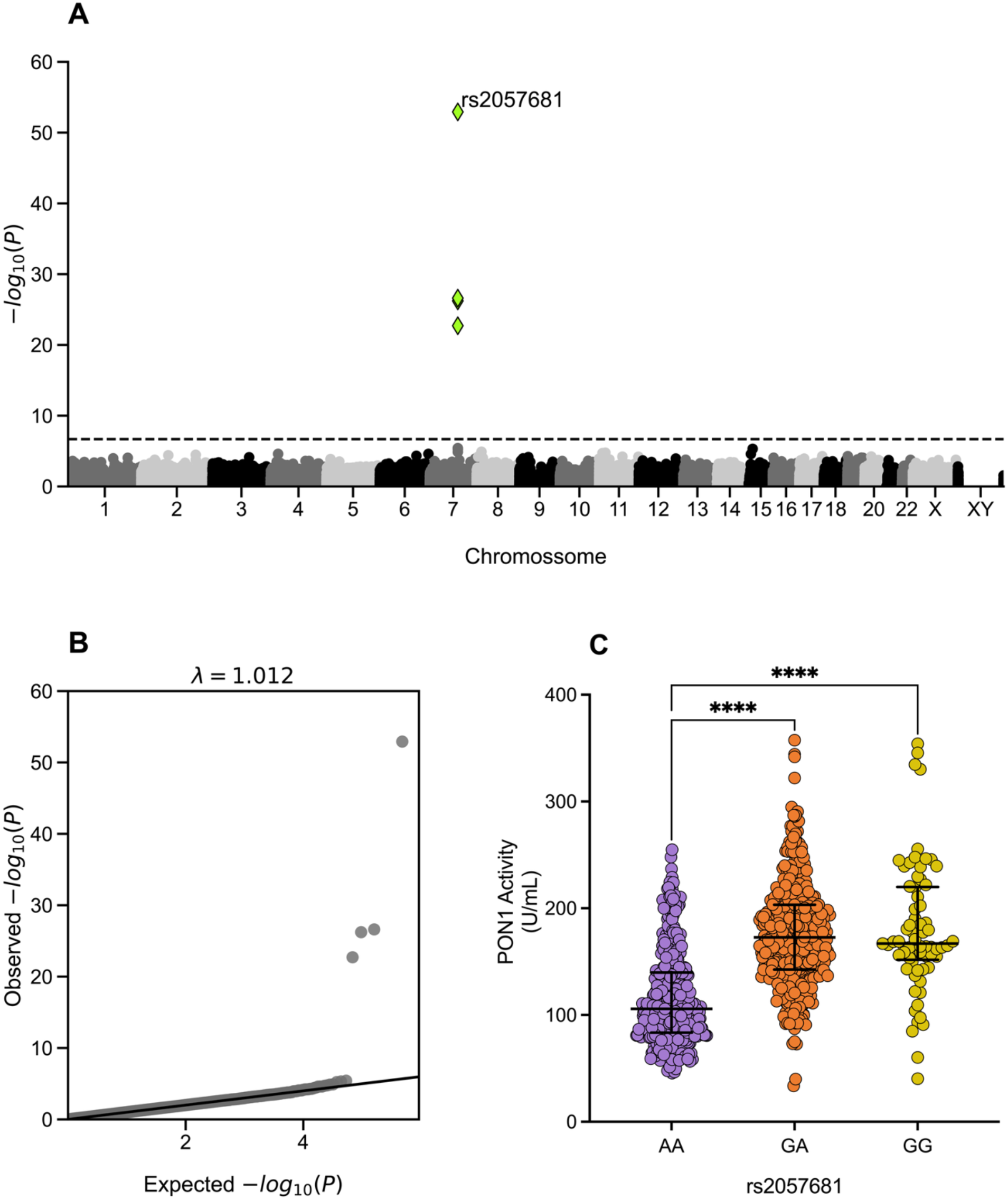
Genome-wide analysis of serum PONase activity in the PREVADIAB2 cohort. A. Manhattan plot showing genome-wide association analysis with Serum PONase activity in the PREVADIAB2 cohort (N = 922). A total of 240,830 SNPs were tested using a linear additive model adjusted for age, sex, and BMI. The x-axis represents genomic position by chromosome, and the y-axis shows –log_10_(P values) of association. The horizontal dashed line represents our GWAS significance threshold (P = 2.07E10-7). The seven SNPs with significant results are marked as green diamonds. The association results peaked at rs2057681 (Padj = 3.99E-48), located in the PON1 gene region in chromosome 7. B. Quantile-Quantile (QQ) plot of the genome-wide association analysis showing observed versus expected -log_10_(P values) under the null hypothesis. The observed inflation factor (λ = 1.012) indicates minimal genomic inflation. The upward deviation at the tail suggests genuine associations.C. Genetic effect of the lead associated SNP, rs2057681, on levels of Serum PONase activity. Genotypes (AA, GA, GG) are shown on the x-axis, and Serum PONase activity on the y-axis. Data points represent individual Serum PONase activity values. Phenotype distributions are overlaid with median and error bars indicating the interquartile range. Genotype groups size: AA = 417, AG = 376, and GG = 69. The G allele is associated with increased Serum PONase activity in a dominant manner. Statistical testing used Kruskal–Wallis test with Dunn’s correction for multiple comparisons. *P < 0.05, ** P < 0.01 and *** P < 0.001.

To assess potential confounding by glycemic status or liver dysmetabolism, a stratified analysis was performed by categorizing participants into normoglycemic or dysglycemic (including prediabetes and diabetes) groups or according to liver fibrosis risk (FNI ≤0.1or FNI >0.1). The genome-wide association analysis results demonstrated that the association between SNPs at the PON1 locus and PONase activity remained robust across strata in both analyses (Supplementary Figure 2 and Supplementary Table 1), indicating that PONase activity is not significantly affected by the presence of dysglycemia or liver fibrosis. This finding is consistent with previous research showing that serum PON1 activity does not differ significantly between individuals with or without diabetes after accounting for genetic and clinical variables. ^35^.

The most significant association with PONase activity was observed at rs2057681, which reached genome-wide significance under both additive (Padj = 3.99 × 10^−48^) and dominant (Padj = 2.04 × 10^−57^) models (Supplementary Table 2). The G allele at rs2057681 was associated with increased PONase activity in a dominant manner (Figure 2C). Notably, rs2057681 is in near-complete LD (r^2^ = 0.99) with rs662 (Q192R), a functional missense variant previously implicated in PON1 enzymatic activity ^36^.

**Figure 2:**
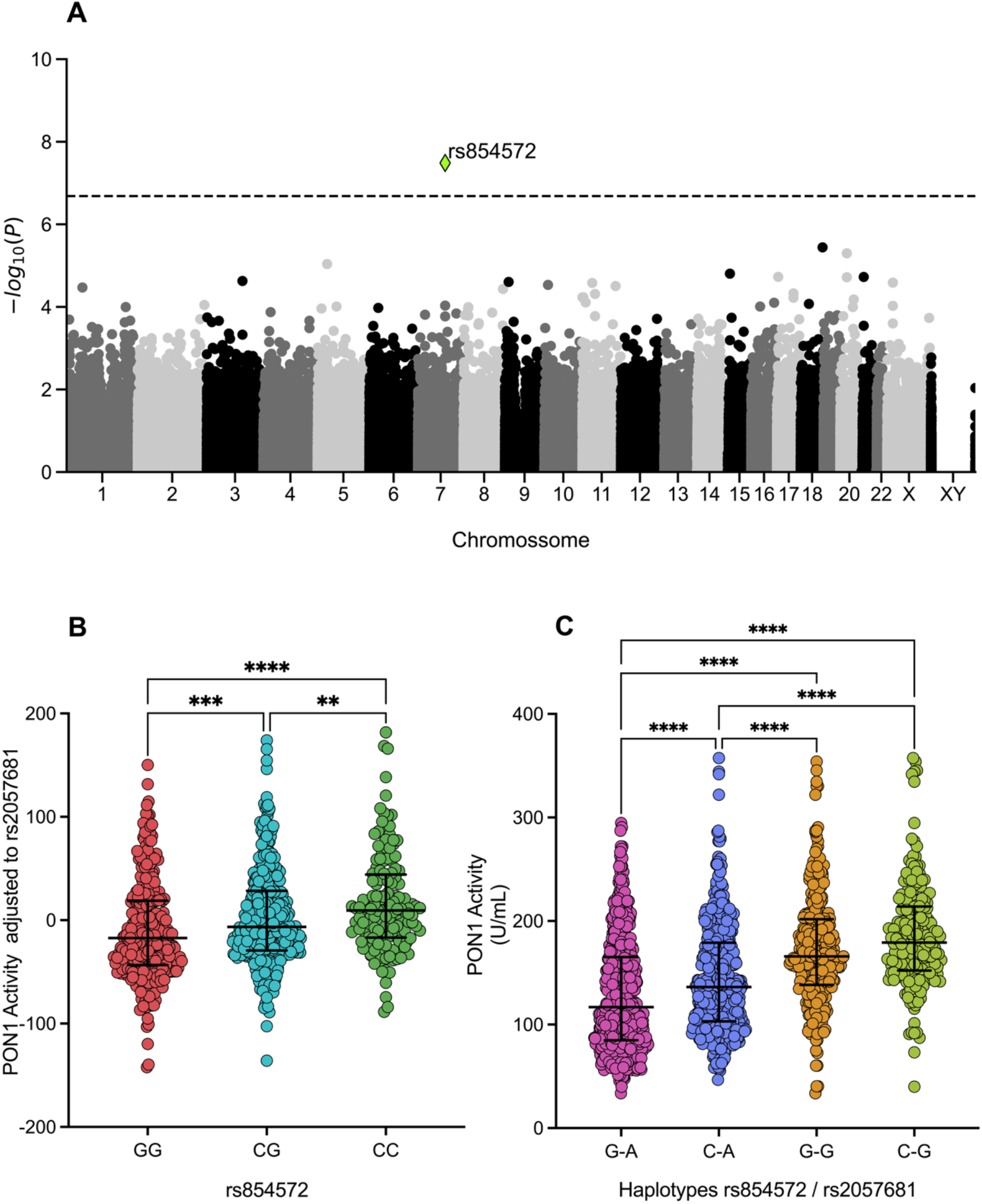
Conditional analysis and combinatorial genetic effects controlling serum PONase activity. A. Manhattan plot showing genome-wide association analysis with Serum PONase activity in the PREVADIAB2 cohort after conditioning on the lead SNP rs2057681. The analysis was performed in 922 individuals with 240,830 SNPs tested using a linear additive model. The x-axis shows chromosomal position, and the y-axis shows –log_10_(P values) of association. The horizontal line marks the genome-wide significance threshold (P = 2,07E-7). After conditioning, rs854572 is the only SNP showing –log10(P values) above the significance threshold. B. Serum PONase activity of rs854572 genotypes (GG, CG, CC) residuals after adjustment for the rs2057681genotype. Violin plots with overlaid box plots show the distribution, median, and interquartile range of PONase activity residuals. Genotype groups size: GG = 313, CG = 399, and CC = 150. C. Association of PON1 haplotypes with serum PONase activity. Haplotypes were constructed from rs854572 and rs2057681 genotypes, resulting in four observed allelic combinations: G–A, C–A, G–G, and C–G (first allele = rs854572, second allele = rs2057681). Violin plots with overlaid box plots show the distribution, median, and interquartile range of PONase activity for each haplotype group. Haplotype groups size: G–A = 705, C–A = 506, G–G = 322, C–G = 193. Statistical testing in B and C used Kruskal–Wallis test with Dunn’s correction for multiple comparisons. *P < 0.05, ** P < 0.01 and *** P < 0.001.

Conditional analysis adjusting for rs2057681 identified rs854572, located in the PON1 promoter region, as an independent genome-wide significant signal (P = 3.19 × 10^−8^; Padj = 9.53 × 10^−3^). The C allele at rs854572 exhibited an additive effect, increasing serum PONase activity (Figure 2 and Supplementary Table 2). This promoter variant, also known as −909 C/G, is not in LD with rs2057681 and resides in a neighboring LD block; it has been previously associated with both PONase and arylesterase activities ^37^. These results indicate that rs854572 exerts a regulatory effect on PON1 gene expression, while rs2057681, located in the structural gene, exerts a distinct effect on enzymatic activity, likely reflecting the Q192R polymorphism.

### PON1 genotypic combinations control PONase activity and dysmetabolic conditions

To investigate how independent PON1 genetic signals jointly influence serum PONase activity and dysmetabolic phenotypes, we focused on two variants identified by genome-wide and conditional analyses: rs2057681, a coding-region variant in near-complete linkage disequilibrium with Q192R, and rs854572, a promoter-region variant exerting an independent regulatory effect on PON1 activity.

Based on phased genotypes, four rs854572–rs2057681 haplotypes were observed: G–A, C– A, G–G, and C–G (first allele rs854572, second allele rs2057681). Comparison of serum PONase activity across haplotypes revealed a graded effect consistent with the individual SNP associations. The C–G haplotype was associated with the highest PONase activity, whereas the G–A haplotype showed the lowest activity. The G–G haplotype conferred higher activity than the C–A haplotype, indicating that the rs2057681 genotype exerts a stronger influence on PONase activity than the promoter variant rs854572 (Figure 2C).

To assess associations between PON1 genotypic combinations and dysmetabolic outcomes, analyses of dysglycemia and liver fibrosis were restricted to individuals aged over 55 years, a subgroup in which these conditions are more prevalent and clinically established. Given the dominant effect of rs2057681 on PONase activity, associations were evaluated after stratification by rs2057681 genotype to determine whether the effect of the promoter variant rs854572 differed according to the coding-region genetic background.

Among individuals carrying at least one G allele at rs2057681 (GA or GG genotypes), the G– A haplotype was associated with the highest prevalence of both dysglycemia and liver fibrosis and was therefore used as the reference group. Within this genetic background, carriers of the C–A haplotype exhibited a significantly lower risk of dysglycemia and liver fibrosis compared with G–A carriers (Tables 2 and 3; Figure 3). In contrast, the G–G and C– G haplotypes did not confer consistent protection relative to G–A, despite being associated with higher PONase activity.

**Table 2:**
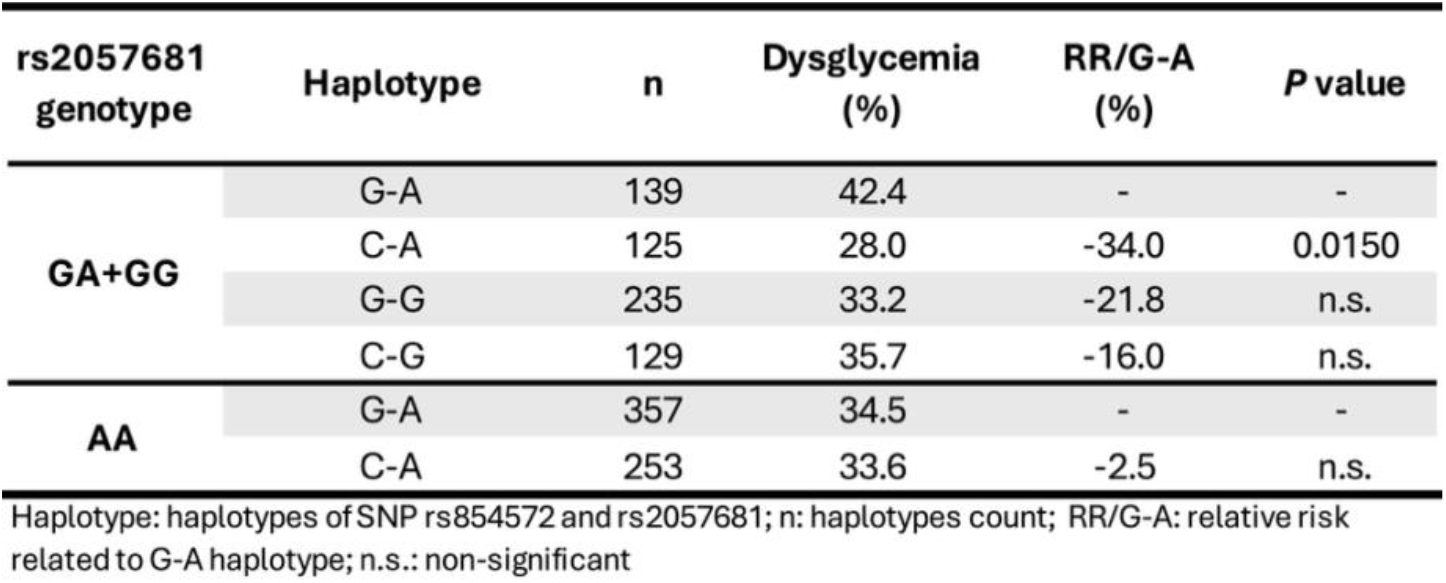
Proportion and relative risk of dysglycemia in PON1 haplotypes carriers stratified by rs2057681 genotype.

**Table 3:** Proportion and relative risk of liver fibrosis in PON1 haplotypes carriers stratified by rs2057681 genotype.

| rs2057681 genotype | Haplotype | n | FNI (>0.1) (%) | RR/G-A (%) | P value |
| --- | --- | --- | --- | --- | --- |
| GA+GG | G-A | 141 | 47.5 | - | - |
|  | C-A | 126 | 33.3 | -29.9 | 0.0246 |
|  | G-G | 237 | 43.9 | -7.7 | n.s. |
|  | C-G | 132 | 46.2 | -2.7 | n.s. |
| AA | G-A | 364 | 43.7 | - | - |
|  | C-A | 258 | 52.3 | 19.8 | 0.0345 |

Haplotype: haplotypes of SNP rs854572 and rs2057681; n: haplotypes count; FNI: Fibrotic NASH Index; RR/G-A: relative risk related to G-A haplotype, n.s.; non-significant

**Figure 3:**
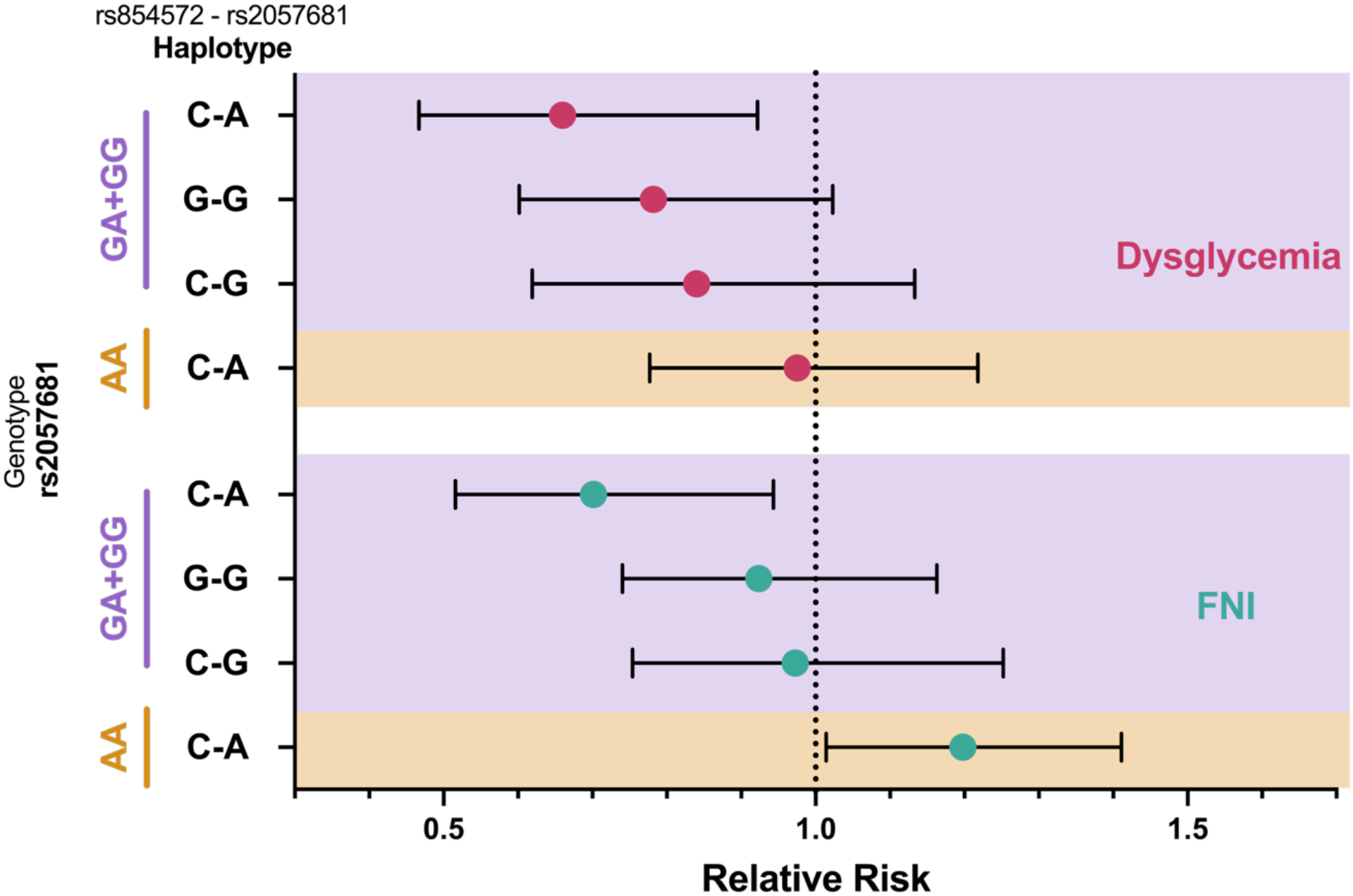
PON1 promoter–coding region haplotypic combinations governing risk of dysglycemia and liver fibrosis. Relative risk of dysglycemia (red) and liver fibrosis (FNI > 0.1; green) conferred by rs854572 (promoter)–rs2057681 (coding region) haplotypic combinations (C–A, G–G, and C–G) relative to the G–A haplotype. Relative risks were estimated in individuals aged over 55 years and stratified by rs2057681 genotype (GA, GG (purple) or AA (orange). Error bars represent 95% confidence intervals.

In individuals homozygous for the A allele at rs2057681 (AA genotype), a distinct pattern was observed. In this context, the C–A haplotype was no longer protective and was instead associated with an increased risk of liver fibrosis compared with the G–A haplotype, while no significant protection against dysglycemia was observed. Thus, the same promoter–coding region combination (C–A) exerted opposite effects on liver fibrosis risk depending on the rs2057681 genotype background.

Collectively, these results demonstrate that PON1 promoter–coding region combinations modulate dysglycemia and liver fibrosis risk in a genotype-dependent manner. While rs854572–rs2057681 haplotypes strongly influence serum PONase activity, disease associations are not explained by enzyme activity alone, but rather by specific genetic configurations defined by both haplotypic structure and coding-region genotype context.

## Discussion

In this study, we show that two independent genetic variants in the PON1 locus—rs2057681 and rs854572—jointly shape serum PONase activity and modulate susceptibility to dysmetabolic conditions in a genotype-dependent manner. While rs2057681, a coding-region variant in near-complete linkage disequilibrium with the functional Q192R polymorphism, emerged as the primary determinant of PONase activity ^38^, rs854572 exerted an independent regulatory effect consistent with a promoter-associated mechanism. Importantly, our results demonstrate that specific promoter–coding region combinations are associated with dysglycemia and liver fibrosis risk in individuals aged over 55 years, independently of their effect on steady-state serum PONase activity.

Consistent with previous reports, rs2057681 showed the strongest association with serum PONase activity, likely reflecting the well-established functional impact of the Q192R polymorphism. The 192R isoform is known to display enhanced hydrolysis of paraoxon but reduced efficiency toward lipid peroxides and other physiological substrates ^39^, which may differentially influence oxidative stress handling depending on metabolic context. The 192R allele frequency varies significantly by ethnicity: it is highest in East Asians (>60%), intermediate in South Asians (∼40%), and lowest in Europeans (<35%). Interestingly, in Asians the 192R allele in disease association it is associated with increased risk for type 2 diabetes and coronary artery disease, while in Europeans, it may be protective or neutral ^40^. In sum, the genetic effects of Q192R alleles on PON1 enzymatic activity and disease association are variable possibly reflecting the intervening role of other genetic factors.

Conditional analysis identified rs854572 as an independent contributor to PONase activity, supporting a role for promoter-region variants in regulating PON1 expression. Although rs854572 itself has limited direct functional annotation, it is in strong linkage disequilibrium with -108C/T (rs705379) and -162A/G (rs705381) polymorphisms that were reported to affect PON1 gene expression and serum enzyme levels ^41^. Prior studies have reported context-dependent associations between rs854572 alleles and lipid traits, PON1 activity, and vascular outcomes, reinforcing the notion that promoter variants exert effects that depend on genetic and environmental context rather than acting uniformly across populations ^42, 43^.

A central finding of our study is that the association between PON1 genetic variation and dysmetabolic outcomes is not explained by PONase activity alone. Although rs854572– rs2057681 haplotypes showed a graded association with serum PONase activity, dysglycemia and liver fibrosis risk followed a distinct pattern that depended on the rs2057681 genotype background. In individuals aged over 55 years carrying at least one rs2057681 G allele, the C–A haplotype was associated with reduced risk of dysglycemia and liver fibrosis relative to the G–A haplotype. In contrast, among rs2057681 AA homozygotes, the same C–A haplotype was associated with increased liver fibrosis risk, illustrating a reversal of effect depending on coding-region genotype context. These findings underscore the importance of genetic context in shaping disease susceptibility. Rather than acting through a simple linear relationship between enzyme activity and disease risk, PON1 promoter and coding-region variants appear to interact in cis (haplotypes) and in trans (genotype classes) to define functional states of the PON1 system. This may involve differences in transcriptional regulation, enzyme stability, substrate specificity, or responsiveness to oxidative and inflammatory challenges that are not captured by basal PONase activity measurements.

The restriction of dysglycemia and liver fibrosis analyses to individuals over 55 years strengthens the clinical relevance of our findings, as metabolic and fibrotic manifestations become more prevalent and stable with aging. This approach reduces heterogeneity related to early or transient metabolic alterations and suggests that PON1 genetic configurations may be particularly informative for identifying susceptibility to clinically meaningful dysmetabolic outcomes in older individuals.

Our results also help reconcile inconsistencies in the literature regarding associations between PON1 variants and metabolic diseases. Numerous studies have reported population-specific, phenotype-specific, or even contradictory effects of individual PON1 polymorphisms ^40,44−46^. By demonstrating that disease associations depend on specific promoter–coding region combinations and genotype backgrounds, our findings provide a genetic framework that may explain these discrepancies and highlight the limitations of single-variant analyses.

Several limitations should be acknowledged. We focused on serum PONase activity as a functional readout, but other PON1 activities, such as arylesterase or lactonase activity, as well as circulating PON1 protein levels, may capture additional disease-relevant mechanisms. Moreover, while our cohort is well characterized, replication in independent populations will be necessary to confirm these findings and to clarify the underlying biological mechanisms.

In conclusion, this study demonstrates that specific PON1 promoter–coding region genetic configurations, rather than serum PONase activity alone, are associated with susceptibility to dysglycemia and liver fibrosis in older adults. These findings highlight the complex genetic architecture underlying PON1 function and support the use of combinatorial genetic profiling to improve risk stratification for dysmetabolic conditions. Future studies exploring how these genetic configurations interact with environmental and inflammatory stressors may further elucidate the role of PON1 in metabolic disease pathogenesis.

## Data Availability

All data presented in this manuscript will be available upon request to collaborative academic researchers for non-profit research purposes only.

## Notes

**Funding statement:** this work was supported by “Fundação para a Ciência e a Tecnologia” – FCT to MJM (PD/BD/114256/2016), LH (2024.03785.BDANA), and the European Commission, Horizon Europe Framework Programme, PAS GRAS (Grant agreement n. 101080329).

**Conflict of interest disclosure:** All the authors declare that they have no conflict of interest.

### Competing Interest Statement

The authors have declared no competing interest.

### Funding Statement

this work was supported by the Fundacao para a Ciencia e a Tecnologia to MJM (PD/BD/114256/2016) and LH (2024.03785.BDANA), and the European Commission, Horizon Europe Framework Programme, PAS GRAS (Grant agreement n. 101080329).

### Author Declarations

The protocol followed the principles of the Declaration of Helsinki and received approval from the Ethics Committee of Associacao Protectora dos Diabeticos de Portugal (APDP Diabetes Portugal (900/2013)) and NOVA Medical School (CEFCM/02/2020), as well as from the Portuguese Data Protection Authority (permit no. 3228/2013).

