## Supplementary information for "Dissecting PON1 Genotype Combinations Modulating Paraoxonase Activity and Risk of Dysglycemia and Liver Fibrosis"

##### Supplementary Figure 1

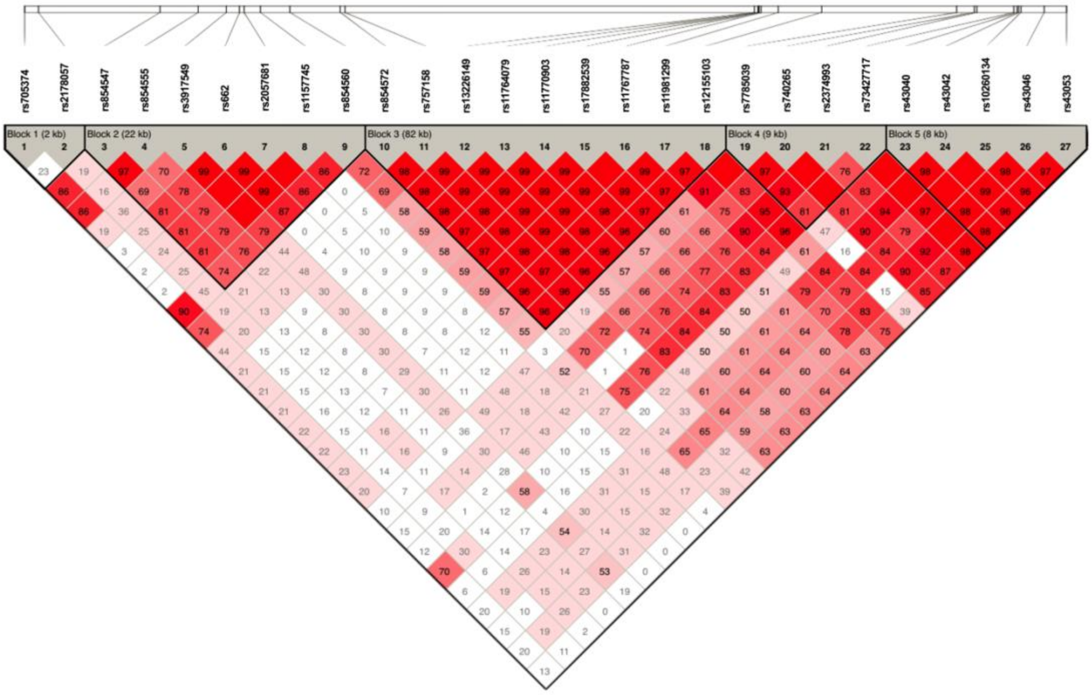


**Supplementary Figure 1:** Linkage Disequilibrium (LD) map of PON1, PON2 and PON3 genes region, generated by the Haploview software (version 4.1). The LD map was constructed with genotype data of 27 SNPs in the chromosome 7 region (95271366 to 95449748 bp) obtained from 922 individuals of the PREVADIAB2 cohort. Relative SNPs positions in chromosome 7 are depicted in upper bar and respective SNP ID are shown. Pair-wise D' values are shown inside each diamond. The strength of LD is color-coded by LOD score, with red diamonds indicating high LD and white diamonds indicating low LD. The black triangles show 5 LD-blocks defined using the Solid Spine of LD method, with the spine extended when D′ > 0.74. The seven SNPs associated to PONase activity in the GWAS are within LD block 2, while rs854572 identified in the conditional analysis is in LD block 3.

**Supplementary Figure 2**

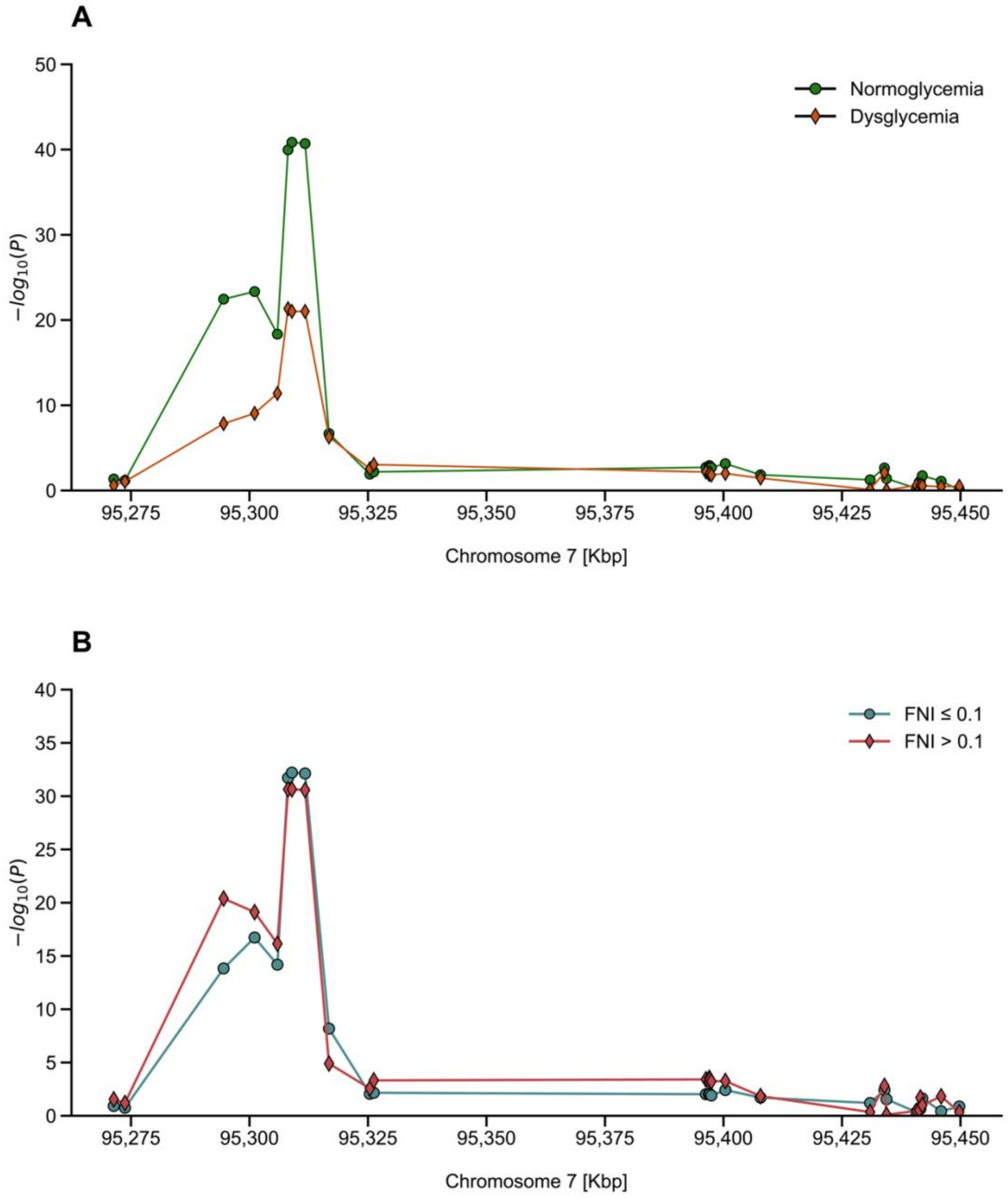


**Supplementary Figure 2:** Association peaks of PONase activity is not affected by glycemic states or liver dysmetabolism. Stratified association analysis of serum PONase activity in the PON genes region. PREVADIAB2 subjects were stratified in (A) normoglycemic (n= 562 in green) and dysglycemic groups (n= 242 in orange) or, (B) according the Fibrosis NASH Index, in FNI ≤.01 (n= 485 in blue) or >0.1 (n= 335 in red). The plots shows results of association association tests (-$\log_{10}$ of *P*-value) and the genomic position of tested SNPs encompassing the PON1, PON2 and PON3 genes region on chromosome 7 (95,271,366–95,449,748 bp).

### Supplementary Table 1

**Supplementary Table 1:** Association of PONase activity to SNPs in the PON1, PON2 and PON3 genes region, before and after stratification for glycemic status (normoglycemia and dysglycemia) or liver fibrosis risk (FNI≤0.1 and FNI>0.1).

| | | | | All Individuals | | | | | | | Normoglycemia | | | Dysglycemia | | | FNI ≤ 0.1 | | | FNI > 0.1 | | |
|---|---|---|---|---|---|---|---|---|---|---|---|---|---|---|---|---|---|---|---|---|---|---|
| CHR | BP | Ref. SNP | Ref. All. | n | HWE | MAF | BETA | STAT | *P* | *P* Adjusted | n | *P* | *P* Adjusted | n | *P* | *P* Adjusted | n | *P* | *P* Adjusted | n | *P* | *P* Adjusted |
| 7 | 95271366 | rs705374 | G | 824 | 0.40 | 0.11 | 11.57 | 2.37 | 1.80E-02 | 4.87E-01 | 561 | 4.48E-02 | 1.00E+00 | 242 | 2.52E-01 | 1.00E+00 | 485 | 1.21E-01 | 1.00E+00 | 334 | 2.87E-02 | 7.74E-01 |
| 7 | 95273727 | rs2178057 | T | 822 | 0.21 | 0.49 | 11.97 | 2.65 | 8.34E-03 | 2.25E-01 | 560 | 6.69E-02 | 1.00E+00 | 242 | 9.02E-02 | 1.00E+00 | 482 | 1.78E-01 | 1.00E+00 | 335 | 6.34E-02 | 1.00E+00 |
| 7 | 95294544 | rs854547 | G | 824 | 0.73 | 0.38 | 45.18 | 12.13 | 3.03E-31 | 8.17E-30 | 562 | 3.54E-23 | 9.55E-22 | 241 | 1.44E-08 | 3.88E-07 | 485 | 1.48E-14 | 3.99E-13 | 334 | 4.03E-21 | 1.09E-19 |
| 7 | 95301079 | rs854555 | A | 825 | 0.78 | 0.37 | 46.39 | 12.57 | 2.83E-33 | 7.63E-32 | 562 | 4.59E-24 | 1.24E-22 | 242 | 8.93E-10 | 2.41E-08 | 485 | 1.86E-17 | 5.01E-16 | 335 | 7.72E-20 | 2.09E-18 |
| 7 | 95305887 | rs3917549 | T | 825 | 0.50 | 0.18 | 46.15 | 11.87 | 4.30E-30 | 1.16E-28 | 562 | 4.54E-19 | 1.23E-17 | 242 | 4.08E-12 | 1.10E-10 | 485 | 6.49E-15 | 1.75E-13 | 335 | 7.14E-17 | 1.93E-15 |
| 7 | 95308134 | rs662 | C | 824 | 0.64 | 0.30 | 60.82 | 18.18 | 2.59E-62 | 6.99E-61 | 561 | 1.08E-40 | 2.91E-39 | 242 | 4.61E-22 | 1.24E-20 | 484 | 1.92E-32 | 5.19E-31 | 335 | 2.27E-31 | 6.14E-30 |
| 7 | 95308945 | rs2057681 | G | 825 | 0.76 | 0.30 | 61.06 | 18.28 | 6.97E-63 | 1.88E-61 | 562 | 1.39E-41 | 3.76E-40 | 242 | 9.76E-22 | 2.64E-20 | 485 | 6.18E-33 | 1.67E-31 | 335 | 2.27E-31 | 6.14E-30 |
| 7 | 95311726 | rs1157745 | T | 823 | 0.64 | 0.30 | 61.13 | 18.26 | 9.73E-63 | 2.63E-61 | 560 | 1.99E-41 | 5.37E-40 | 242 | 9.76E-22 | 2.64E-20 | 484 | 7.43E-33 | 2.01E-31 | 334 | 2.65E-31 | 7.17E-30 |
| 7 | 95316772 | rs854560 | T | 824 | 1.00 | 0.39 | -29.50 | -7.41 | 3.08E-13 | 8.32E-12 | 561 | 2.09E-07 | 5.63E-06 | 242 | 5.17E-07 | 1.40E-05 | 485 | 6.56E-09 | 1.77E-07 | 334 | 1.26E-05 | 3.41E-04 |
| 7 | 95325384 | rs854572 | C | 824 | 0.30 | 0.40 | 16.01 | 3.92 | 9.56E-05 | 2.58E-03 | 561 | 1.21E-02 | 3.26E-01 | 242 | 2.82E-03 | 7.61E-02 | 484 | 8.65E-03 | 2.33E-01 | 335 | 2.70E-03 | 7.30E-02 |
| 7 | 95326216 | rs757158 | T | 825 | 0.40 | 0.38 | 17.60 | 4.40 | 1.22E-05 | 3.29E-04 | 562 | 6.58E-03 | 1.78E-01 | 242 | 9.10E-04 | 2.46E-02 | 485 | 6.95E-03 | 1.88E-01 | 335 | 4.74E-04 | 1.28E-02 |
| 7 | 95396288 | rs13226149 | A | 824 | 0.64 | 0.23 | 17.12 | 4.29 | 1.99E-05 | 5.38E-04 | 562 | 1.93E-03 | 5.21E-02 | 241 | 6.68E-03 | 1.80E-01 | 485 | 9.46E-03 | 2.55E-01 | 334 | 3.85E-04 | 1.04E-02 |
| 7 | 95396917 | rs11764079 | T | 825 | 0.71 | 0.23 | 16.69 | 4.19 | 3.13E-05 | 8.44E-04 | 562 | 1.93E-03 | 5.21E-02 | 242 | 1.21E-02 | 3.27E-01 | 485 | 9.46E-03 | 2.55E-01 | 335 | 6.23E-04 | 1.68E-02 |
| 7 | 95397015 | rs11770903 | G | 825 | 0.71 | 0.23 | 17.18 | 4.32 | 1.79E-05 | 4.82E-04 | 562 | 1.18E-03 | 3.17E-02 | 242 | 1.21E-02 | 3.27E-01 | 485 | 9.46E-03 | 2.55E-01 | 335 | 2.81E-04 | 7.59E-03 |
| 7 | 95397096 | rs17882539 | A | 825 | 0.71 | 0.23 | 17.09 | 4.29 | 1.98E-05 | 5.35E-04 | 562 | 1.86E-03 | 5.02E-02 | 242 | 9.19E-03 | 2.48E-01 | 485 | 9.19E-03 | 2.48E-01 | 335 | 3.69E-04 | 9.96E-03 |
| 7 | 95397441 | rs11767787 | C | 822 | 0.85 | 0.23 | 16.50 | 4.13 | 3.96E-05 | 1.07E-03 | 561 | 2.02E-03 | 5.44E-02 | 240 | 1.51E-02 | 4.08E-01 | 484 | 1.24E-02 | 3.35E-01 | 333 | 5.95E-04 | 1.61E-02 |
| 7 | 95400389 | rs11981299 | A | 819 | 1.00 | 0.21 | 17.75 | 4.43 | 1.06E-05 | 2.86E-04 | 557 | 7.01E-04 | 1.89E-02 | 241 | 1.01E-02 | 2.72E-01 | 481 | 3.86E-03 | 1.04E-01 | 333 | 5.45E-04 | 1.47E-02 |
| 7 | 95407819 | rs12155103 | A | 822 | 0.84 | 0.20 | 13.91 | 3.39 | 7.32E-04 | 1.98E-02 | 561 | 1.45E-02 | 3.90E-01 | 240 | 3.55E-02 | 9.59E-01 | 484 | 1.97E-02 | 5.33E-01 | 333 | 1.43E-02 | 3.85E-01 |
| 7 | 95430930 | rs7785039 | T | 824 | 0.93 | 0.23 | -7.74 | -1.92 | 5.53E-02 | 1.00E+00 | 562 | 5.64E-02 | 1.00E+00 | 241 | 7.74E-01 | 1.00E+00 | 485 | 6.31E-02 | 1.00E+00 | 334 | 4.46E-01 | 1.00E+00 |
| 7 | 95433958 | rs740265 | G | 819 | 0.41 | 0.23 | 16.49 | 4.15 | 3.62E-05 | 9.77E-04 | 558 | 2.27E-03 | 6.13E-02 | 240 | 7.82E-03 | 2.11E-01 | 482 | 3.94E-03 | 1.06E-01 | 332 | 1.58E-03 | 4.27E-02 |
| 7 | 95434390 | rs2374993 | G | 822 | 0.90 | 0.16 | -8.09 | -1.87 | 6.25E-02 | 1.00E+00 | 560 | 3.99E-02 | 1.00E+00 | 241 | 9.35E-01 | 1.00E+00 | 483 | 2.81E-02 | 7.60E-01 | 334 | 7.97E-01 | 1.00E+00 |
| 7 | 95440708 | rs73427717 | A | 822 | 0.91 | 0.18 | -0.42 | -0.10 | 9.22E-01 | 1.00E+00 | 559 | 5.68E-01 | 1.00E+00 | 242 | 2.45E-01 | 1.00E+00 | 484 | 4.53E-01 | 1.00E+00 | 333 | 3.47E-01 | 1.00E+00 |
| 7 | 95441264 | rs43040 | C | 822 | 0.57 | 0.23 | -6.34 | -1.57 | 1.17E-01 | 1.00E+00 | 560 | 1.12E-01 | 1.00E+00 | 241 | 7.21E-01 | 1.00E+00 | 484 | 2.08E-01 | 1.00E+00 | 333 | 2.94E-01 | 1.00E+00 |
| 7 | 95441538 | rs43042 | A | 824 | 0.65 | 0.32 | -9.19 | -2.32 | 2.07E-02 | 5.57E-01 | 561 | 7.24E-02 | 1.00E+00 | 242 | 2.29E-01 | 1.00E+00 | 485 | 2.25E-01 | 1.00E+00 | 334 | 1.95E-02 | 5.25E-01 |
| 7 | 95441941 | rs10260134 | T | 821 | 0.86 | 0.25 | 11.13 | 2.81 | 5.01E-03 | 1.35E-01 | 560 | 1.84E-02 | 4.97E-01 | 240 | 2.74E-01 | 1.00E+00 | 482 | 2.34E-02 | 6.32E-01 | 334 | 1.01E-01 | 1.00E+00 |
| 7 | 95445918 | rs43046 | A | 821 | 0.88 | 0.33 | -8.51 | -2.14 | 3.25E-02 | 8.78E-01 | 559 | 8.07E-02 | 1.00E+00 | 241 | 3.65E-01 | 1.00E+00 | 484 | 3.75E-01 | 1.00E+00 | 332 | 1.50E-02 | 4.06E-01 |
| 7 | 95449748 | rs43053 | C | 818 | 0.68 | 0.41 | -3.09 | -0.74 | 4.57E-01 | 1.00E+00 | 558 | 8.36E-01 | 1.00E+00 | 239 | 3.17E-01 | 1.00E+00 | 481 | 1.35E-01 | 1.00E+00 | 332 | 4.51E-01 | 1.00E+00 |

Results are based on dominant genetic model adjusted for age, sex, and BMI. CHR: chromosome; BP: base pair position (GRCh38,p14); Ref. SNP: reference SNP ID; Ref. All.: reference allele; n: number of subjects; HWE: Hardy–Weinberg equilibrium p-value; MAF: minor allele frequency; BETA: regression coefficient; STAT: t-statistic; *P*: nominal p-value; *P* Adjusted: Bonferroni-adjusted p-value for the full cohort; FNI: Fibrotic NASH Index.

### Supplementary Table 2

**Supplementary Table 2:** Association of PONase activity to SNPs in the PON1, PON2 and PON3 genes region using an additive genetic model before and after conditioning for rs2057681.

| CHR | BP | Ref. SNP | Ref. All. | n | HWE | MAF | STAT | Additive Model *P* | Additive Model *P* Adj. | Conditional Model *P* | Conditional Model *P* Adj. |
|---|---|---|---|---|---|---|---|---|---|---|---|
| 7 | 95271366 | rs705374 | G | 824 | 0.40 | 0.11 | 2.60 | 9.45E-03 | 2.55E-01 | 1.18E-02 | 2.84E-01 |
| 7 | 95273727 | rs2178057 | T | 822 | 0.21 | 0.49 | 3.14 | 1.76E-03 | 4.76E-02 | 3.46E-01 | 1.00E+00 |
| 7 | 95294544 | rs854547 | G | 824 | 0.73 | 0.38 | 11.14 | 6.00E-27 | 1.62E-25 | 4.63E-01 | 1.00E+00 |
| 7 | 95301079 | rs854555 | A | 825 | 0.78 | 0.37 | 11.77 | 1.19E-29 | 3.22E-28 | 1.47E-01 | 1.00E+00 |
| 7 | 95305887 | rs3917549 | T | 825 | 0.50 | 0.18 | 11.23 | 2.46E-27 | 6.64E-26 | 3.69E-01 | 1.00E+00 |
| 7 | 95308134 | rs662 | C | 824 | 0.64 | 0.30 | 16.50 | 5.14E-53 | 1.39E-51 | | |
| 7 | 95308945 | rs2057681 | G | 825 | 0.76 | 0.30 | 16.61 | 1.20E-53 | 3.23E-52 | | |
| 7 | 95311726 | rs1157745 | T | 823 | 0.64 | 0.30 | 16.63 | 1.03E-53 | 2.77E-52 | | |
| 7 | 95316772 | rs854560 | T | 824 | 1.00 | 0.39 | -10.29 | 1.96E-23 | 5.29E-22 | 9.32E-05 | 2.24E-03 |
| 7 | 95325384 | rs854572 | C | 824 | 0.30 | 0.40 | 4.65 | 3.83E-06 | 1.04E-04 | 3.27E-08 | 7.84E-07 |
| 7 | 95326216 | rs757158 | T | 825 | 0.40 | 0.38 | 5.00 | 6.97E-07 | 1.88E-05 | 7.72E-08 | 1.85E-06 |
| 7 | 95396288 | rs13226149 | A | 824 | 0.64 | 0.23 | 4.60 | 4.92E-06 | 1.33E-04 | 1.34E-04 | 3.22E-03 |
| 7 | 95396917 | rs11764079 | T | 825 | 0.71 | 0.23 | 4.36 | 1.47E-05 | 3.98E-04 | 2.15E-04 | 5.15E-03 |
| 7 | 95397015 | rs11770903 | G | 825 | 0.71 | 0.23 | 4.38 | 1.36E-05 | 3.68E-04 | 1.70E-04 | 4.08E-03 |
| 7 | 95397096 | rs17882539 | A | 825 | 0.71 | 0.23 | 4.36 | 1.44E-05 | 3.89E-04 | 2.25E-04 | 5.40E-03 |
| 7 | 95397441 | rs11767787 | C | 822 | 0.85 | 0.23 | 4.19 | 3.06E-05 | 8.26E-04 | 2.51E-04 | 6.03E-03 |
| 7 | 95400389 | rs11981299 | A | 819 | 1.00 | 0.21 | 4.68 | 3.44E-06 | 9.30E-05 | 1.81E-04 | 4.34E-03 |
| 7 | 95407819 | rs12155103 | A | 822 | 0.84 | 0.20 | 3.39 | 7.39E-04 | 2.00E-02 | 1.11E-02 | 2.66E-01 |
| 7 | 95430930 | rs7785039 | T | 824 | 0.93 | 0.23 | -2.34 | 1.96E-02 | 5.30E-01 | 8.06E-01 | 1.00E+00 |
| 7 | 95433958 | rs740265 | G | 819 | 0.41 | 0.23 | 4.56 | 5.99E-06 | 1.62E-04 | 5.71E-03 | 1.37E-01 |
| 7 | 95434390 | rs2374993 | G | 822 | 0.90 | 0.16 | -2.21 | 2.72E-02 | 7.35E-01 | 3.93E-01 | 1.00E+00 |
| 7 | 95440708 | rs73427717 | A | 822 | 0.91 | 0.18 | 0.22 | 8.27E-01 | 1.00E+00 | 3.72E-01 | 1.00E+00 |
| 7 | 95441264 | rs43040 | C | 822 | 0.57 | 0.23 | -1.64 | 1.01E-01 | 1.00E+00 | 1.33E-01 | 1.00E+00 |
| 7 | 95441538 | rs43042 | A | 824 | 0.65 | 0.32 | -2.78 | 5.53E-03 | 1.49E-01 | 4.73E-01 | 1.00E+00 |
| 7 | 95441941 | rs10260134 | T | 821 | 0.86 | 0.25 | 3.43 | 6.35E-04 | 1.71E-02 | 3.66E-02 | 8.79E-01 |
| 7 | 95445918 | rs43046 | A | 821 | 0.88 | 0.33 | -2.70 | 7.07E-03 | 1.91E-01 | 5.70E-01 | 1.00E+00 |
| 7 | 95449748 | rs43053 | C | 818 | 0.68 | 0.41 | -1.13 | 2.57E-01 | 1.00E+00 | 9.58E-02 | 1.00E+00 |

Results obtained after adjustment for age, sex, and BMI. CHR: chromosome; BP: base pair position (GRCh38 p14); Ref. SNP: reference SNP ID; Ref. All.: reference allele; n: number of subjects; HWE: Hardy–Weinberg equilibrium p-value; MAF: minor allele frequency; STAT: t-statistic from the allelic model; *P*: nominal p-value; *P* Adj.: Bonferroni-adjusted p-value.
